# faers: A High-Fidelity Framework and R/Bioconductor Package for Precision Adverse Event Surveillance

**DOI:** 10.64898/2026.03.26.26349444

**Authors:** Zhangyu Wang, Yun Peng, Jian-Guo Zhou, Xiaoyun Bu, Yankun Zhao, Zimo Li, Bin Yan, Yujun Sun, Chunyang Wang, Chenyang Shu, Yanru Cui, Shixiang Wang

## Abstract

**Background:** The FDA Adverse Event Reporting System (FAERS) is a critical pillar of post-marketing pharmacovigilance; however, its utility is constrained by data heterogeneity, pervasive reporting redundancies, and inconsistent medical terminology. These structural barriers impede reproducible, large-scale analyses and the implementation of precision drug safety surveillance.

**Methods:** We developed faers, an open-source R package that delivers a standardized framework and an end-to-end workflow for transforming raw FAERS data into analysis-ready formats. The package implements a regulatory-compliant multi-level deduplication strategy, automated MedDRA terminology mapping, and an R S4-based object-oriented system to ensure data integrity, traceability, and efficient management of complex relational structures. It further integrates a full suite of disproportionality signal detection methods, including the Reporting Odds Ratio (ROR), Proportional Reporting Ratio (PRR), Bayesian Confidence Propagation Neural Network (BCPNN), and Empirical Bayes Geometric Mean (EBGM). Performance was benchmarked on large-scale FAERS datasets, and validity was confirmed by replicating published findings on anti-PD-1/PD-L1-associated cardiotoxicity and CAR-T cell therapy outcomes, with additional application to immune-related adverse events (irAEs).

**Findings:** The package demonstrated high computational efficiency and near-linear scalability when processing extensive quarterly FAERS data. Validation analyses of two case studies showed excellent concordance with prior literature. Application to an irAE cohort further identified a statistically significant age-by-sex interaction in risk patterns, demonstrating the tool’s ability to uncover nuanced demographic signals that are often missed by conventional approaches.

**Interpretation:** The faers package provides a transparent, scalable, and fully reproducible framework for FAERS-based pharmacovigilance. By automating data cleaning, standardization, and advanced signal detection, it lowers technical barriers for researchers and regulators while promoting high-quality, open pharmacoepidemiological research to strengthen drug safety monitoring.

**Research in Context:** *Evidence before this study:* Existing pharmacovigilance resources, ranging from R packages like PhViD and openEBGM to web-based platforms such as openFDA, have significantly advanced statistical signal detection. However, these tools primarily focus on specific algorithms and require pre-processed, high-quality input data, leaving a critical gap in end-to-end data management. No comprehensive, open-source framework currently integrates the entire workflow—from raw FAERS data ingestion and regulatory-compliant deduplication to advanced signal detection—within a single, reproducible environment. Consequently, large-scale studies often rely on fragmented, custom scripts that hinder scalability and research transparency.

*Added value of this study:* We developed faers, an integrated, high-fidelity R package that streamlines the pharmacovigilance workflow by unifying regulatory-compliant data preprocessing—including multi-level deduplication and MedDRA mapping—with a comprehensive suite of frequentist and Bayesian signal detection methods. Benchmarking demonstrated near-linear scalability on large-scale datasets, and the tool’s analytical accuracy was validated through the successful replication of representative studies on ICI-associated cardiotoxicity and CAR-T therapy outcomes. Furthermore, an exploratory analysis of immune-related adverse events revealed a significant age-by-sex interaction in reporting risk, illustrating the package’s capacity to uncover nuanced demographic patterns that are often obscured by conventional analytical approaches.

*Implications of all the available evidence:* The R/Bioconductor package faers establishes a standardized, scalable, and fully reproducible framework for FAERS-based pharmacovigilance. By automating the transition from raw, heterogeneous data to actionable signal detection, it lowers technical barriers for researchers, clinicians, and regulators alike. This advancement promotes greater transparency and rigor in pharmacoepidemiology, ultimately strengthening post-marketing drug safety surveillance and providing a robust foundation for precision clinical practice and regulatory decision-making.

*Availability:* The faers package is freely available through the Bioconductor repository at https://bioconductor.org/packages/faers/. Its source code is openly hosted on GitHub at https://github.com/WangLabCSU/faers.

## 1. Introduction

With the rapid expansion of the global pharmaceutical market and the emergence of innovative therapies, post-marketing safety surveillance has become a cornerstone of public health. Traditional pharmacovigilance systems were primarily optimized for Type A adverse drug reactions (ADRs)—those that are dose-dependent and predictable—associated with conventional small-molecule drugs^1^. However, these systems often falter when confronted with the complex, mechanism-based, and immune-mediated toxicities characteristic of modern biological therapies, which frequently deviate from traditional dose-response paradigms^2^. In this context, the advent of precision medicine has not only catalyzed a fundamental paradigm shift in drug development but has also necessitated a transition in safety monitoring from detecting dose-dependent non-specific toxicity to the precise identification and prediction of mechanism-driven specific toxicity^3^.

Immune-modulating agents, exemplified by immune checkpoint inhibitors (ICIs), have revolutionized oncology by reshaping immune homeostasis to exert anti-tumor effects^4^. However, this unique biological mechanism has also introduced a distinct toxicity profile, named immune-related adverse events (irAEs), which differs significantly from traditional chemotherapy. The severity of irAEs lies not only in their broad organ involvement and clinical heterogeneity (affecting tissues from the skin and gastrointestinal tract to the heart and nervous system) but also in their unpredictable onset and rapid progression^5^. Certain severe irAEs, such as immune-related myocarditis and pneumonitis, present narrow therapeutic windows and high fatality risks. Notably, the case fatality rate for immune-related myocarditis has been reported as high as 46%^6^, and the incidence of fatal irAEs in combination immunotherapy can reach 1.23%^7^.

Despite this, clinical risk assessment for irAEs remains heavily reliant on pre-marketing randomized controlled trials (RCTs)^8^. While RCTs are the gold standard for evaluating drug efficacy, their inherent methodological limitations hinder the capture of the full spectrum of toxicity in real-world settings. First, stringent inclusion criteria systematically exclude complex clinical populations, such as the elderly, patients with multiple comorbidities, or those with a history of autoimmune diseases, who are at the highest risk for irAEs in clinical practice^9^. Second, limited sample sizes (typically hundreds to thousands) are insufficient to detect rare but fatal toxicities with incidences below 1%^10^. Finally, the relatively short follow-up periods in RCTs fail to reveal the long-term evolutionary patterns of late-onset irAEs^11^. This evidence gap leaves clinicians without precise risk-stratification tools for real-world patients. Consequently, there is an urgent clinical need to leverage large-scale monitoring data to map the landscape of irAEs, identify rare immune toxicities, and characterize their temporal dynamics to optimize immunotherapy decision-making and ensure patient safety.

The FDA Adverse Event Reporting System (FAERS), maintained by the U.S. Food and Drug Administration, is one of the world’s largest and most authoritative databases for spontaneous ADR reporting. By aggregating vast data from healthcare professionals, patients, and manufacturers, FAERS serves as a critical resource for identifying and evaluating potential drug safety signals, and its public availability has significantly advanced pharmacovigilance research^12^. However, the complex relational structure of FAERS, characterized by multi-table associations, inconsistent data formats across reporting periods, and pervasive information redundancy (e.g., duplicate reports). This poses significant technical challenges for extracting actionable insights^13–15^. The lack of a standardized preprocessing framework often leads to substantial methodological heterogeneity, undermining the transparency and comparability of pharmacovigilance evidence. Furthermore, existing platforms (e.g., FAERS Public Dashboard^16^ and openFDA^17^) primarily offer basic query functionalities and operate as black-box systems with limited programmatic flexibility. These tools fail to meet the rigorous requirements for large-scale epidemiological modeling, such as data cleaning, customized deduplication, and workflow automation, nor can they efficiently process high-dimensional historical data spanning decades. Given that R has become the cornerstone of modern statistical computing and epidemiology, there remains a notable absence of a systematic, reproducible analysis toolkit specifically designed for the R environment that integrates memory-efficient data structures with administrative-grade analytical workflows^18^.

To address these gaps, we developed faers, an R/Bioconductor package designed as a high-fidelity pipeline for precision adverse event surveillance. This toolkit provides an end-to-end, scriptable framework that streamlines the entire pharmacovigilance workflow, from raw data acquisition and rigorous preprocessing to signal detection. By integrating MedDRA-based terminology mapping with robust disproportionality analysis, faers ensures high data fidelity and computational scalability. By unifying complex drug safety workflows within the R environment, this package not only enhances the reproducibility and transparency of large-scale epidemiological studies but also provides a robust technical foundation for precision pharmacovigilance and evidence-based drug safety monitoring.

## 2. Results

### 2.1 Design Philosophy and Overall Architecture

The faers R package is designed to bridge the gap between raw regulatory data and the modern bioinformatics ecosystem (**Figure 1**). Embracing an end-to-end pharmacovigilance workflow, the package integrates data acquisition, processing, and analysis within the R environment, facilitating the seamless transformation of raw case records into actionable scientific evidence. Architecturally, we adopted an object-oriented paradigm—utilizing R’s S4 class system—to abstract FAERS data by encapsulating quarterly submissions into unified container objects. By embedding metadata, such as standardization status and deduplication logs, directly into these objects, the package ensures the transparency and traceability of the analytical process. The core FAERS class maintains data integrity through a slot-based mechanism that tracks file formats (ASCII/XML), temporal dimensions, and processing states, thereby converting heterogeneous raw data into semantically consistent, structured objects. To balance out-of-the-box usability with the flexibility required for customized analysis, we employed a modular design pattern, implementing each stage of the data pipeline as an independent, composable functional unit. A master controller function orchestrates these modules into an automated pipeline while allowing researchers to intervene at any stage; this design not only ensures the reproducibility of the scientific workflow but also provides an extensible interface for complex, cross-quarter analyses. Through parsing standardization, the system utilizes concurrent downloads and integrity checks to parse heterogeneous ASCII and XML files into standardized S4 objects, automatically managing field mapping and enriching temporal metadata to provide a unified interface for data since 2004. 2) The ***Clinical Terminology Standardization*** module harmonizes records via the MedDRA hierarchy, employing a two-tier matching mechanism (from lowest level terms to preferred terms) and integrating the Athena drug knowledge base for bulk drug name normalization and synonym resolution, thereby providing a semantic foundation for theme-based analysis. 3) Subsequently, the ***Muti-level Deduplication Strategy*** module employs a rule-based algorithm that executes record linkage across eight critical dimensions—including sex, age, reporting country, event date, therapy start date, indication, drug name, and adverse event. The system utilizes a six-round iterative strategy that enforces exact matching for drug and adverse event fields while allowing tolerance for auxiliary fields, effectively identifying duplicate cases originating from multiple sources while preventing spurious clusters via unique encoding for missing values. 4) Finally, the ***Pharmacovigilance Signal Detection*** module, based on a unified contingency table framework, integrates a comprehensive suite of statistical methods. Frequentist approaches include the Proportional Reporting Ratio (PRR), Reporting Odds Ratio (ROR), and associated statistical tests, while Bayesian methods encompass two implementations of the Bayesian Confidence Propagation Neural Network (BCPNN)—normal approximation and Markov Chain Monte Carlo (MCMC) variants—as well as the Empirical Bayes Geometric Mean (EBGM). The system leverages parallel computing to process drug-event pairs, outputting structured signal lists containing point estimates, confidence intervals, and statistical significance metrics to support clinical evaluation.

**Figure 1.**
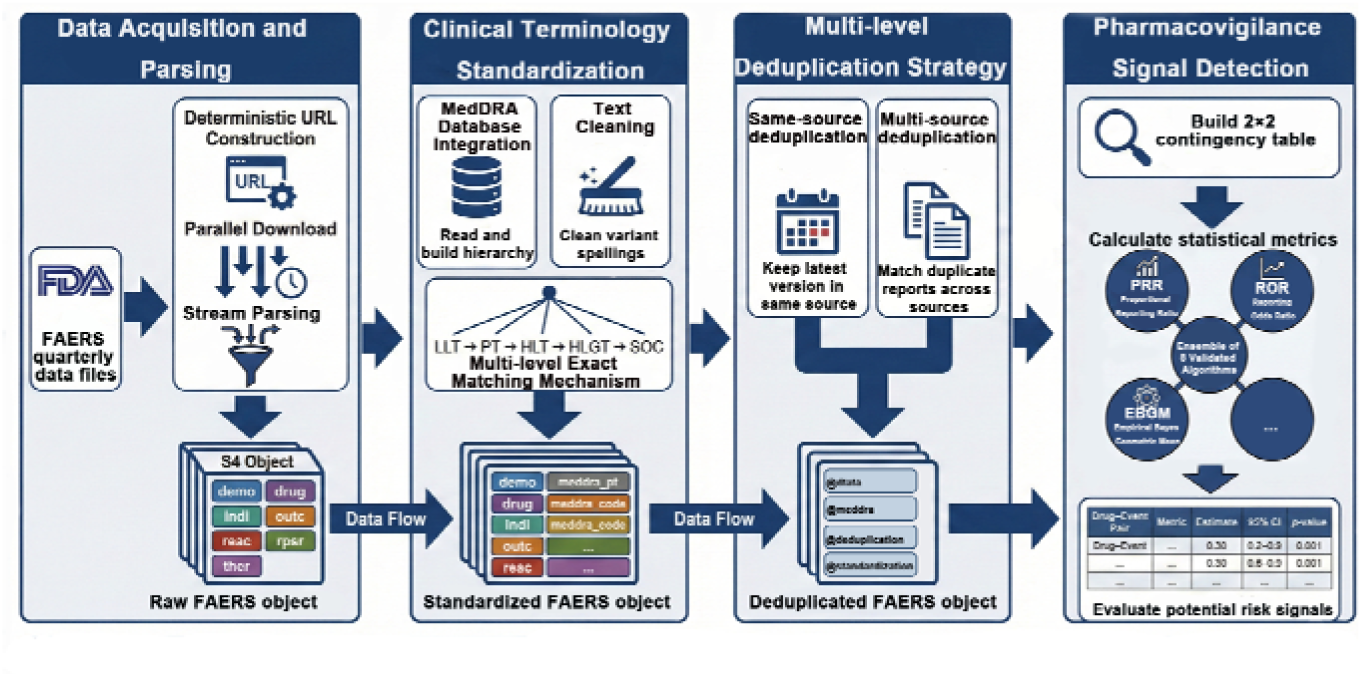
Integrated workflow of the faers package for FAERS data analysis. Building upon these design principles, the framework automates the workflow from raw data ingestion to signal generation through four core functional modules: 1) The ***Data Acquisition and Parsing*** module serves as the system entry point, automatically retrieving raw data from official FDA sources.

Given the computational challenges posed by the massive datasets inherent in pharmacovigilance research, performance optimization was prioritized as a core design objective. By leveraging the R package data.table for memory-efficient data manipulation and integrating parallel computing strategies, faers enables high-performance processing of large-scale datasets. Furthermore, the package offers extensible interfaces to facilitate the future integration of pharmacovigilance data with other high-dimensional data modalities, such as genomic profiles, thereby providing a robust technical foundation for precision pharmacovigilance practices.

### 2.2 Benchmark of Performance and Scalability

To evaluate the computational efficiency of the faers pipeline, we conducted a benchmark using the full 2015 FAERS dataset, which was pre-downloaded to isolate processing time from network latency. The total processing time for this dataset was 2.39 minutes (**Figure 2A**). Modular performance profiling indicated that the multi-layered deduplication stage accounted for 50.2% (1.20 minutes) of the total runtime, while the signal detection module required 3.46 seconds (<3% of the total duration). The peak memory footprint for the entire workflow was 17.26 GB, compared to the cumulative memory requirement of 17.16 GB for the individual functional modules.

**Figure 2.**
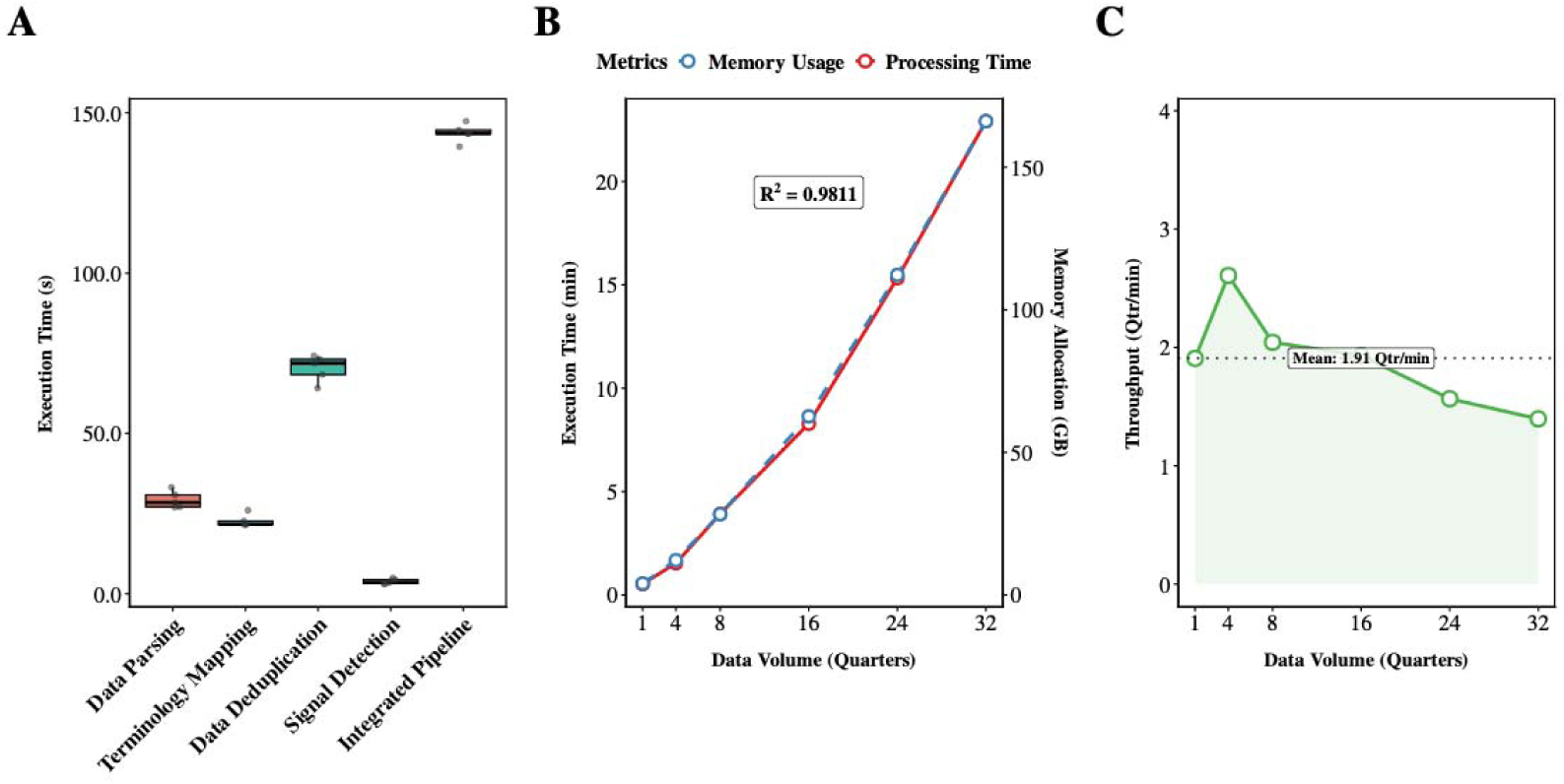
Performance profiling and scalability analysis of the faers workflow. (A) Execution time distribution for core workflow components. Boxplots with jittered points show median execution times across five iterations; duplicate detection exhibits the highest runtime. (B) Scalability analysis. Dual-axis plot showing processing time and peak memory allocation as data volume increases from 1 to 32 quarters, with near-linear growth (R² = 0.9811). (C) Processing efficiency. Area plot showing throughput (quarters per minute) across increasing data volumes; the mean throughput remains consistent at 1.91 quarters per minute.

Scalability stress tests were performed by increasing input volume from one to 32 quarters (eight years of data). Both processing time and memory allocation increased linearly with input volume (**Figure 2B**). Linear regression analysis of processing time yielded an R^2^ of 0.9811. At the maximum scale of 32 quarters, the analysis was completed in 22.5 minutes, with a system throughput of 1.91 quarters per minute (**Figure 2C**).

### 2.3 Case Study

To evaluate the accuracy and robustness of the faers pipeline, we conducted three case studies. The first two studies validated key findings from previous FAERS-based pharmacovigilance research, while the third performed an exploratory analysis of demographic interactions in immune-related adverse events.

#### Case Study I: Replication of PD-1/PD-L1-Associated Severe Cardiac Toxicity

We replicated the study by Cheng et al. (2024) regarding severe cardiac adverse events (scAEs) associated with anti-PD-1/PD-L1 therapies^19^. We adhered to the original study’s data scope, drug and event definitions, and statistical methods. The entire workflow, from data acquisition to signal detection, was executed using the faers package. The analysis yielded results consistent with the original findings across three dimensions: data foundation, core signal strength, and patient population characteristics (**Table 1**).

**Table 1:**
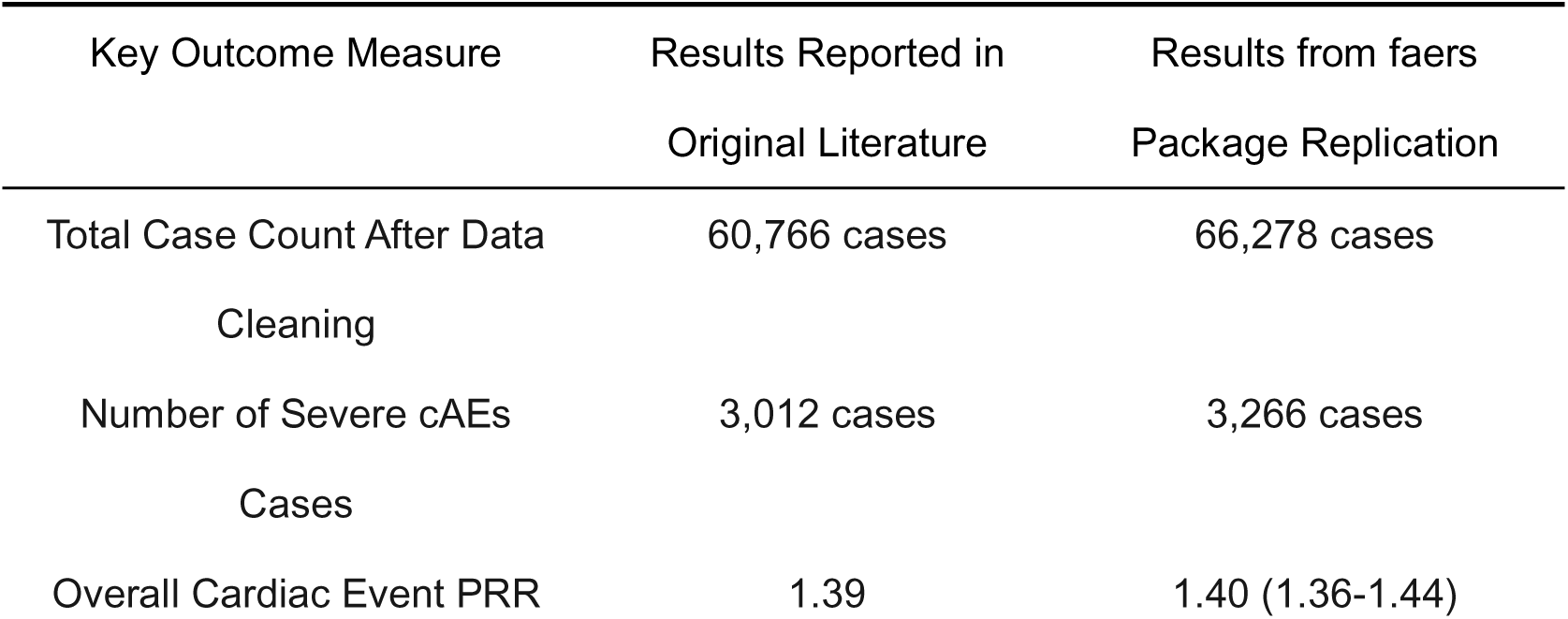
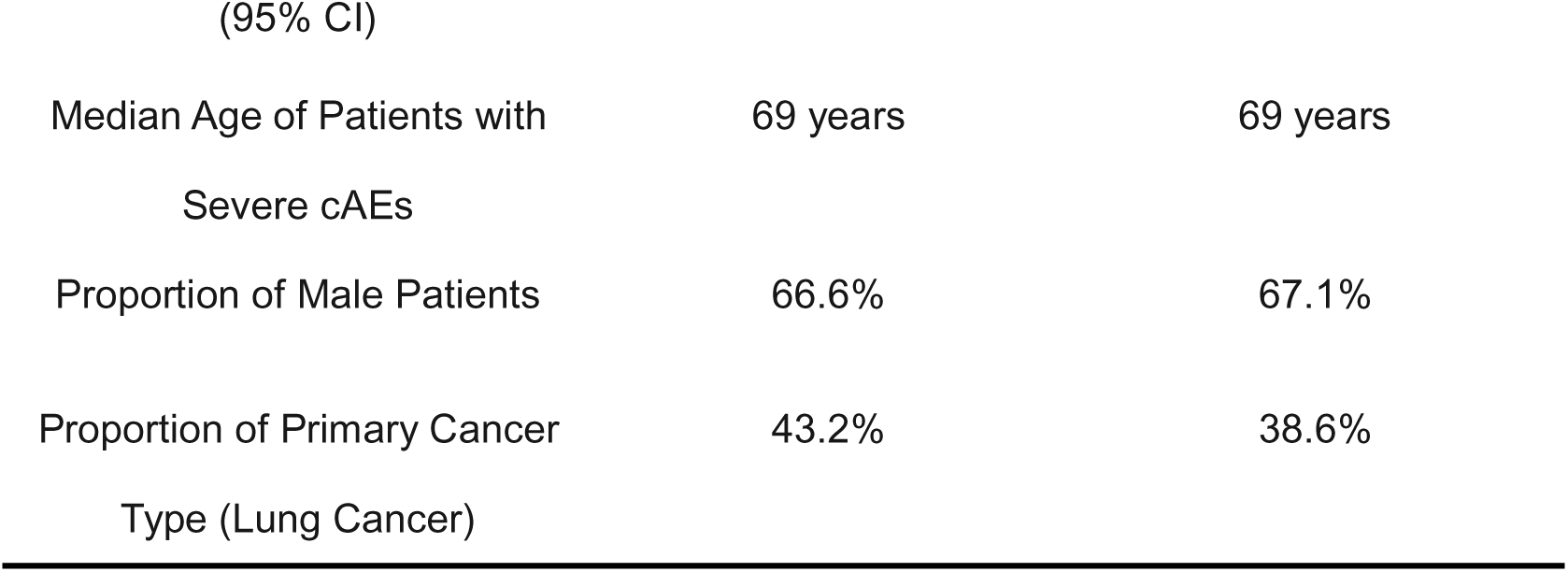
Comparison of Key Results from replication study on Anti-PD-1/PD-L1 cardiotoxicity.

The analysis identified high-intensity signals for ICI-associated cardiac toxicity, as illustrated in the volcano plot (**Figure 3A**), where myocarditis, pericarditis, and heart failure emerged as the top five signals by PRR. Demographic analysis of the scAE cohort revealed a median age of 69 years (**Figure 3B**), with a male predominance (67.1%; **Figure 3C**). Temporal analysis indicated a rapid onset of cardiac toxicity, with a median time-to-onset of 37 days following ICI initiation, and the majority of events occurring within the first 60 days (**Figure 3D**). Furthermore, these events were associated with significant clinical impact, characterized by a high proportion of serious outcomes, including hospitalization and death (**Figure 3E**).

**Figure 3.**
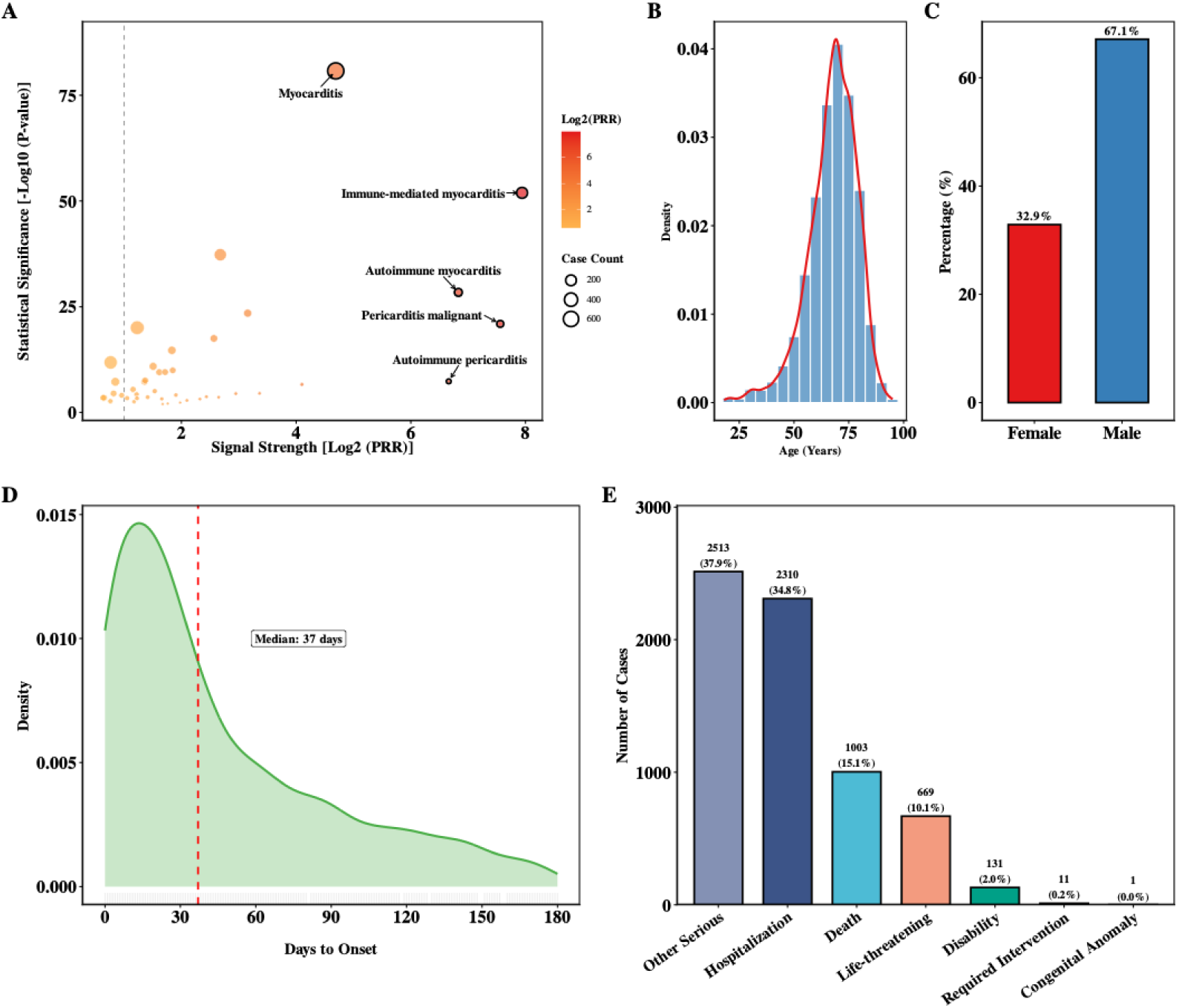
Integrated analysis of ICI-associated scAEs. (A) Volcano plot of disproportionate reporting signals. X-axis denotes signal strength (log_2_(PRR)), and Y-axis represents statistical significance -log_10_(p). Top five signals by PRR are labeled. (B) Age distribution. Density plot showing the age distribution of patients with scAEs. (C) Gender proportion. Bar chart showing the gender distribution of patients with scAEs. (D) Time-to-onset analysis. Kernel density plot showing the time from ICI initiation to cardiac event (median: 37 days). (E) Clinical outcomes. Bar chart showing the distribution of serious outcomes (death, hospitalization, and others) among patients with scAEs.

#### Case Study II: Validation of Secondary Primary Malignancies Following CAR-T Cell Therapy

We replicated the study by Peng et al. (2025) regarding the association between antibiotic exposure and secondary primary malignancies (SPMs) in patients receiving CAR-T cell therapy. We adhered to the original study’s data scope, drug and event definitions, and statistical methods. The entire workflow, from data acquisition to signal detection and interaction analysis, was executed using the faers package. The analysis yielded results highly consistent with the original findings regarding cohort construction, drug identification, and baseline characteristics (**Table 2**).

**Table 2.**
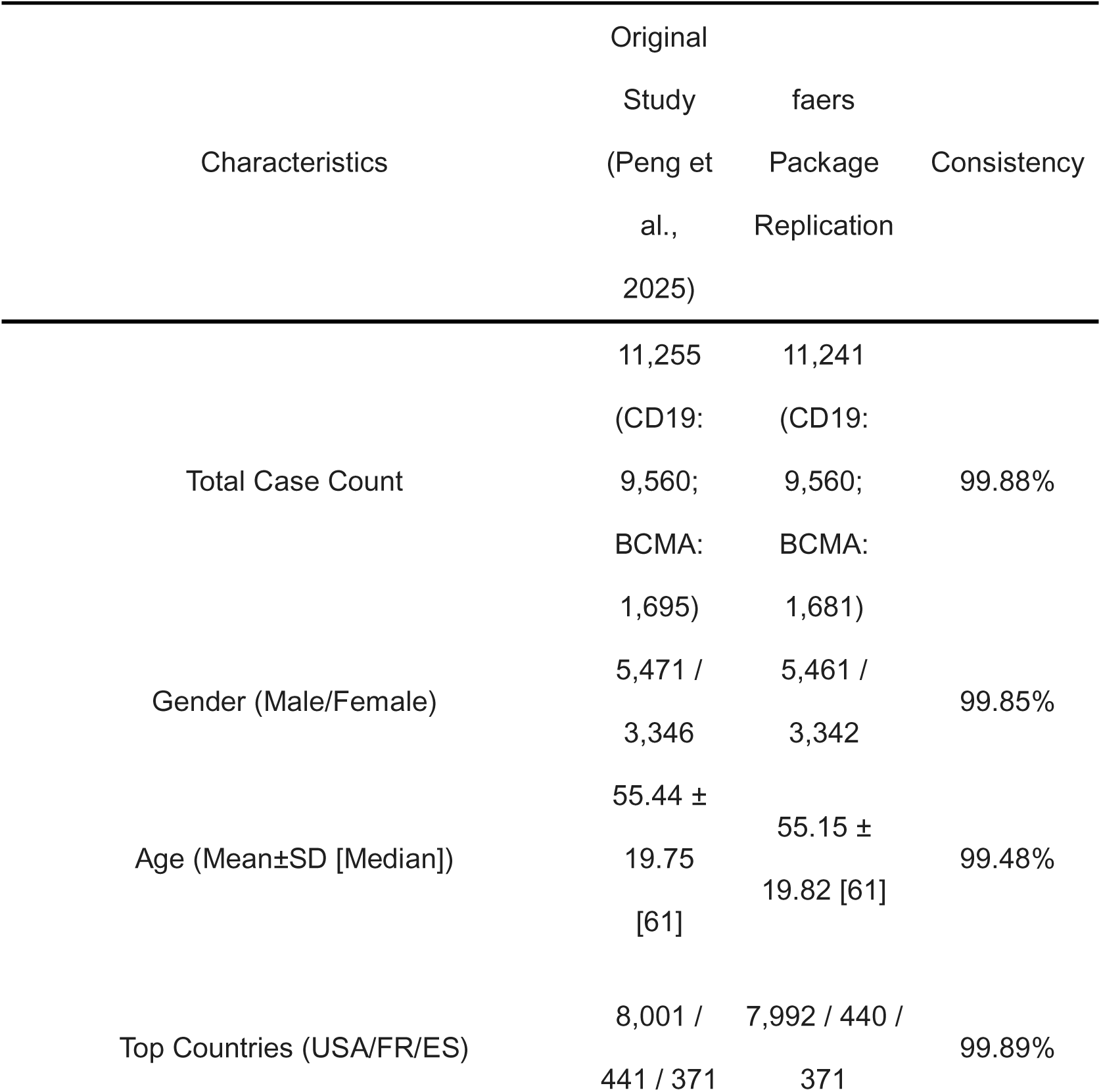
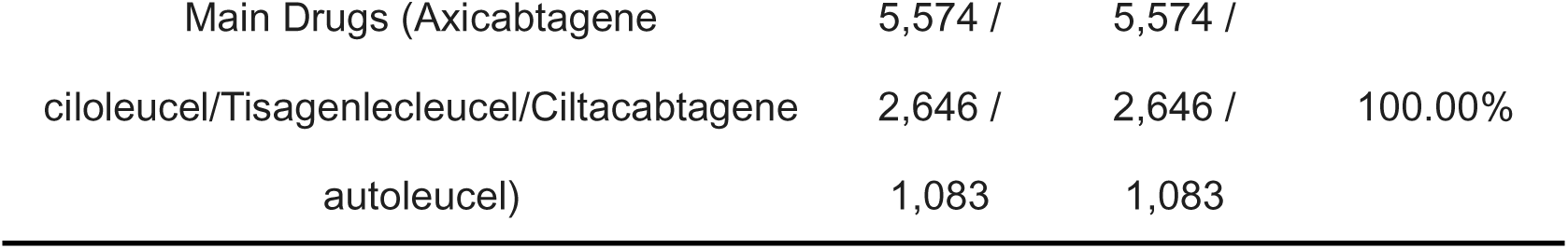
Comparison of baseline characteristics.

Subgroup analysis demonstrated a significant association between antibiotic exposure and SPM risk, as illustrated by the forest plot (**Figure 4A**), where both the overall CAR-T cohort and the CD19-targeted group demonstrated elevated disproportionality signals. Temporal analysis revealed a distinct kinetic pattern of SPM onset; cumulative risk analysis indicated that patients in the antibiotic-exposed group exhibited a significantly higher risk compared to the unexposed group (**Figure 4B**; *p* < 0.0001). This trend remained consistent in the CD19 subgroup (**Figure 4C**; *p* < 0.0001), whereas the BCMA group showed a similar trend that did not reach statistical significance (**Figure 4D**; *p* = 0.075).

**Figure 4.**
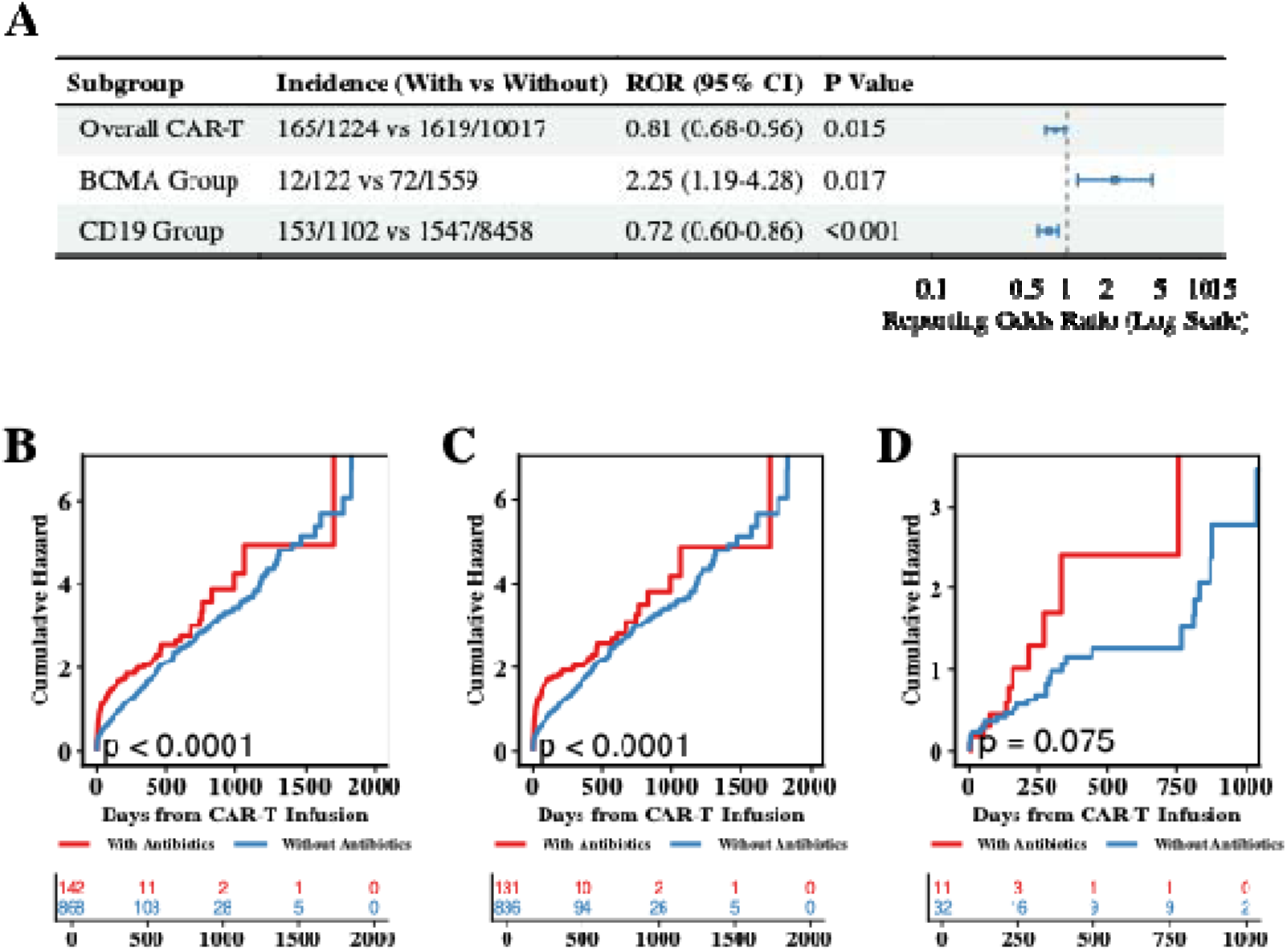
Replication of antibiotic exposure and SPM risk in CAR-T therapy. (A) Forest plot of subgroup analyses. The plot shows the association between antibiotic exposure and SPM risk across the overall CAR-T cohort, the BCMA subgroup, and the CD19 subgroup. (B) Cumulative hazard in overall CAR-T cohort. Time-to-onset analysis comparing patients with and without antibiotic exposure (*p* < 0.0001). (C) Subgroup analysis for CD19. Cumulative hazard comparison between antibiotic-exposed and unexposed patients (*p* < 0.0001). (D) Subgroup analysis for BCMA. Cumulative hazard comparison between antibiotic-exposed and unexposed patients (*p* = 0.075). Below each hazard plot, risk tables show the number of patients at risk over time (days since CAR-T infusion).

#### Case Study III: Interaction Analysis of Sex and Age on the Reporting Risk of Immune-Related Adverse Events

Using the faers package, we extracted reports of ICIs (including PD-1, PD-L1, and CTLA-4 inhibitors) from 2015 to 2025. Descriptive analysis revealed disparities in the proportion and distribution of irAE reports between females and males across four age groups: 18–44, 45–64, 65–74, and ≥75 years (**Figure 5A**). Multivariate logistic regression analysis confirmed that age significantly modulates the effect of gender on the risk of irAE reporting (interaction *p* < 0.001; Likelihood Ratio Test (LRT) = 21.03), with all Generalized Variance Inflation Factors (GVIF) < 2.0. Adjusted predicted probability curves demonstrated that the reporting gap between females and males was most pronounced in the 18–44 age group, narrowing progressively with age, and converging in the ≥75 age group, where 95% confidence intervals overlapped (**Figure 5B**).

**Figure 5.**
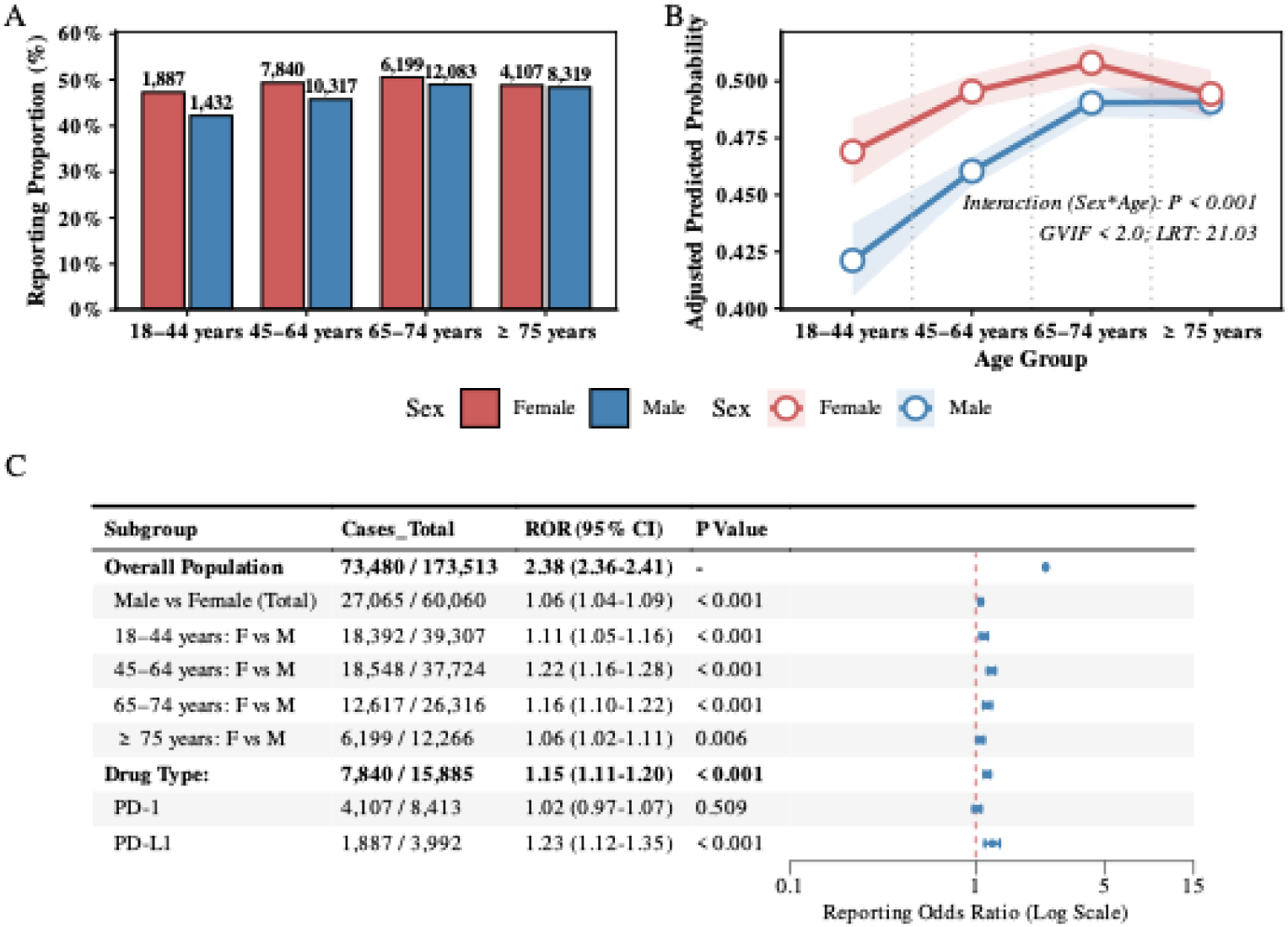
Interaction of sex and age on ICI-associated irAE reporting risk. (A) Observed irAE reporting proportions by age cohort and sex. (B) Adjusted predicted probabilities from multivariable logistic regression. (C) Forest plot of subgroup-specific RORs for demographic and pharmacological factors.

Quantitative analysis of reporting risk revealed significant disparities among ICI users. The ROR for PD-L1 inhibitor users was 1.23 (95% CI: 1.12–1.35; *p* < 0.001), whereas the ROR for PD-1 inhibitor users was 1.02 (95% CI: 0.97–1.07; *p* = 0.509) (**Figure 5C**). Regarding gender disparities stratified by age, the overall ROR for females versus males was 2.38 (95% CI: 2.36–2.41). Stratified analysis indicated an age-dependent attenuation of the gender effect: the ROR for females versus males was 1.11 (95% CI: 1.05–1.16; *p* < 0.001) in the 18–44 age group, 1.22 (95% CI: 1.16–1.28; *p* < 0.001) in the 45–64 group, 1.16 (95% CI: 1.10–1.22; *p* < 0.001) in the 65–74 group, and decreased to 1.06 (95% CI: 1.02–1.11; *p* = 0.006) in the ≥75 age group (**Figure 5C**).

## 3. Discussion

The inherent complexity of FAERS data—characterized by historical format evolution, multi-table associations^20^, and pervasive redundancy^15^—renders it challenging for high-fidelity analysis. While existing web-based platforms (e.g., openFDA^17^) enhance data accessibility, their black-box nature limits programmatic flexibility for complex epidemiological modeling. Conversely, established R packages like PhViD^21^ and openEBGM^22^ provide rigorous statistical frameworks but often require pre-processed inputs, lacking robust capabilities for raw data parsing, iterative deduplication, and terminology standardization. The faers package bridges this gap by providing a transparent, end-to-end workflow that integrates raw data acquisition with advanced analysis. By leveraging an object-oriented S4 framework, faers ensures data integrity through structured metadata management and FDA-aligned iterative deduplication. Furthermore, its rule-based standardization module incorporates nearly 100 normalization rules and MedDRA hierarchy mapping^23^, which transforms unstructured text^24^ into semantically consistent identifiers. Unlike existing tools, this comprehensive preprocessing capability enables researchers to move beyond basic signal detection to sophisticated modeling, such as the multi-confounder adjusted interaction models demonstrated in our case studies. Ultimately, by unifying data management and statistical modeling within a single R environment, faers enhances the reproducibility and comparability of pharmacovigilance evidence, providing a robust technical foundation for precision adverse event surveillance.

The faers package is engineered to handle the massive data volumes inherent in FAERS, demonstrating near-linear scalability in both execution time and memory allocation. Benchmarking on the 2015 dataset revealed a total processing time of 2.39 minutes, with the deduplication stage accounting for 50.2% (1.20 minutes) of the runtime, whereas signal detection required only 3.46 seconds (<3% of the total duration). This performance disparity reflects the distinct computational complexities of these modules: deduplication is a compute-intensive process involving multi-table joins, string aggregation, and iterative fuzzy matching based on FDA rules; in contrast, signal detection relies on lightweight numerical operations on pre-aggregated contingency tables, benefiting significantly from vectorization and parallelization. This predictable performance ensures that scaling from quarterly to decadal analyses does not incur disproportionate computational overhead. Furthermore, by leveraging data.table for memory-efficient manipulation and a modular, stepwise workflow that minimizes intermediate object storage, faers mitigates the out-of-memory risks often associated with large-scale S4 objects. Consequently, the pipeline’s optimized algorithms enable high-volume data processing on standard research workstations, democratizing large-scale drug safety surveillance by removing the need for specialized high-performance computing (HPC) resources.

Through the replication of two representative studies—focusing on PD-1/PD-L1 inhibitor-associated scAEs (Cheng et al.) and antibiotic interactions with SPMs in CAR-T therapy (Peng et al.), we demonstrated the robustness of faers in handling complex clinical data associations. Our results align closely with previous findings, confirming faers’ precision in multi-table association analysis and its ability to reproduce core clinical trends. In the replication of Cheng et al.’s study, we observed a notable discrepancy: while the case count identified by faers was lower (212 vs. 295), the signal strength was markedly higher (PRR = 245.60 vs. 77.01). After accounting for external confounding factors such as drug definitions and data windows, quantitative analysis revealed that this divergence stems primarily from the multi-level deduplication strategy implemented in faers. Spontaneous reporting systems are inherently prone to data inflation due to redundant records. By integrating case records across quarters and multiple sources, faers effectively purifies the denominator, correcting the signal-dilution effect caused by redundant entries. This refinement not only enhances detection sensitivity but also provides a more authentic estimation of drug-event associations, revealing a more robust risk profile.

Furthermore, the sex–age interaction analysis highlights faers’ utility in uncovering biological mechanisms underlying drug safety signals. Our findings indicate a significant sex disparity in irAE risk, with females exhibiting higher reporting rates, potentially reflecting sex-specific immune pathways and hormonal variations^25^. The observed age-dependent attenuation of this gender effect—converging in the ≥75 years group—aligns with previous reports^26^. While these findings are exploratory and limited by inherent reporting biases (e.g., treatment selection bias in older patients), they underscore faers’ capacity for complex stratified analyses, offering a novel perspective for precision, individualized medication assessments in future real-world studies.

Despite the advancements faers offers in data processing and standardization, several limitations warrant consideration. First, as an analytical framework for spontaneous reporting systems, the utility of faers is intrinsically bounded by the quality and nature of the underlying FAERS data. Users must interpret disproportionality signals with caution, acknowledging inherent biases such as the Weber effect, under-reporting, and the inability to establish definitive causality. Second, while the rule-based module effectively maps high-frequency terms, rare or highly complex drug descriptions may still require manual curation. Furthermore, full utilization of the semantic mapping capabilities requires a MedDRA license, which remains a prerequisite for advanced hierarchical analysis.

Future development will prioritize enhancing the tool’s intelligence and multi-source integration capabilities. We aim to incorporate natural language processing (NLP), particularly large language models (LLMs), to extract semantic insights from unstructured narrative text, capturing clinical context often lost in structured fields. Additionally, we plan to integrate faers with broader pharmacological and biological frameworks, such as the Anatomical Therapeutic Chemical (ATC) classification system and proteomics databases. These advancements will facilitate a transition from isolated signal detection to multi-omics-driven association analysis, providing a more mechanistic and precise foundation for real-world drug safety evaluation.

## 4. Conclusion

The faers package provides a high-performance, integrated framework that streamlines the pharmacovigilance workflow, from automated data acquisition to signal detection. By implementing regulatory-grade deduplication and MedDRA-compliant terminology mapping, the package addresses the structural complexities and data redundancy inherent in raw FAERS records. Its accuracy and utility are demonstrated through the successful replication of landmark studies and the discovery of age-dependent sex disparities in irAE risk. In conclusion, faers lowers barriers to large-scale drug safety surveillance, fostering a more standardized and reproducible research ecosystem.

## 5. Methods

### 5.1 Data Sources and Extraction

All raw data from the FAERS were acquired and preprocessed in this study using the faers R package. Primary data were retrieved from the FDA’s official quarterly ASCII files, covering all safety reports from 2004 to 2025. Terminology standardization was performed using the Medical Dictionary for Regulatory Activities (MedDRA, version 26.0).

### 5.2 Benchmarking and Scalability Protocols

The performance of the pharmacovigilance pipeline implemented using the faers package was systematically evaluated with the bench R package^27^. All benchmarks were conducted on a HPC platform equipped with 192 CPU cores and 1.08 TB of RAM. Initial memory states were strictly controlled by forcing garbage collection (gc()) prior to each run.

Baseline Benchmarking: The bench::mark function was used to independently stress-test four core pipeline steps: (1) data parsing (using the faers function with full-year 2015 data); (2) terminology standardization; (3) record deduplication; and (4) signal detection (targeting seven immune checkpoint inhibitors). Each step was executed for five independent iterations to derive median execution time and peak memory allocation. Finally, an end-to-end test of the full workflow was performed to assess overall operational efficiency from raw data ingestion to final output generation.

Scalability Analysis: System scalability with increasing data volume was assessed using the bench::press function. Test loads were configured to simulate data from 1, 4, 8, 16, 24, and 32 quarters, cumulatively spanning from Q1 2014 to Q4 2021. Linear regression models were applied to analyze the relationship between the number of quarters processed and the median execution time, with the coefficient of determination (R²) used to evaluate linear scaling characteristics. Additionally, processing throughput—defined as the number of quarters processed per minute—was calculated to evaluate the operational stability of the system under large-scale workloads.

### 5.3 Experimental Design and Implementation for Case Studies

#### Case Study I: Replication of PD-1/PD-L1-Associated Severe Cardiac Toxicity

To replicate the study by Cheng et al. (2024)^19^, we utilized the faers package to analyze cardiotoxicity associated with anti-PD-1/PD-L1 monotherapy. We adhered to the original study’s data scope (Q1 2014–Q4 2022), drug definitions, and statistical protocols. The entire workflow, from data acquisition to signal detection, was executed using the faers pipeline.

We first applied the faers_filter function to isolate reports where one of seven PD-1/PD-L1 inhibitors—nivolumab, pembrolizumab, cemiplimab, atezolizumab, avelumab, durvalumab, or dostarlimab—was identified as the primary suspect (PS) drug in patients aged ≥18 years. Reports with missing sex or age information were excluded. To rigorously control for potential confounding, we systematically excluded reports involving: (1) cardiac tumors as an indication; (2) concomitant use of known cardiotoxic agents (e.g., doxorubicin, epirubicin, or specific EGFR/ALK inhibitors); or (3) concomitant anti-CTLA-4 therapy.

Subsequently, Preferred Terms (PTs) within the “Cardiac Disorders” System Organ Class (SOC) were extracted based on MedDRA version 26.0. Disproportionality analysis was performed using the faers_phv_signal function. Serious cardiac adverse events were defined as signals meeting two criteria: a minimum of three reported cases and a lower bound of the 95% confidence interval (CI) for the PRR exceeding 1. Finally, time-to-onset distributions were calculated by linking the therapy and demographic tables via unique case identifiers.

#### Case Study II: Validation of Secondary Primary Malignancies Following CAR-T Cell Therapy

In accordance with the protocol established by Peng et al. (2025)^28^, this study assessed the association between antibiotic exposure and the risk of SPMs using over 11,000 reports of CAR-T cell therapy from the second quarter of 2017 to the first quarter of 2024. Cases in which anti-CD19 or anti-BCMA therapies were designated as the primary suspect drugs were identified through regex-based matching and stratified into antibiotic-exposed and non-exposed groups based on concomitant medication records. SPM cases were identified using the MedDRA version 26.0 dictionary (SOC 10029104). Statistical significance was evaluated with Fisher’s exact test, and the ROR along with its 95% confidence interval was calculated. Furthermore, by integrating drug, therapy, and demographic data to extract key temporal endpoints, we employed the Nelson-Aalen cumulative hazard model to analyze differences in time-to-onset (TTO) kinetics between the two groups.

#### Case Study III: Interaction Analysis of Sex and Age on the Reporting Risk of Immune-Related Adverse Events

This study investigated the potential interactions between sex and age on the reporting risk of irAEs associated with ICIs, including PD-1, PD-L1, and CTLA-4 inhibitors, using FAERS data from 2015 to 2025. We utilized the faers package to identify reports where these agents were listed as primary suspect (PS) drugs. The study population was stratified by sex and four age groups (18–44, 45–64, 65–74, and ≥75 years). irAEs were standardized using predefined PTs mapped via the MedDRA hierarchy.

Statistical analyses involved calculating ROR for each age-sex subgroup. To assess the synergistic effects of sex and age on irAE reporting probability, we fitted a multivariable logistic regression model incorporating sex, age, and their interaction term, while adjusting for drug class (PD-1/PD-L1 inhibitors vs. CTLA-4 inhibitors). The significance of the interaction term was evaluated using the LRT.

## Data Availability

All data produced in the present work are contained in the manuscript

https://github.com/MadDERt/faers-reproducibility

## Contributors

ZW, YP and JGZ contributed equally to this work. ZW, YP, and SW were responsible for software development and methodology. ZW, YP, GJZ, and SW contributed to the conceptualization and study design. ZW, YP, JGZ, XB, YZ, ZL, BY, YS, CW and CS performed data curation, formal analysis and visualization. SW, YP, GJZ, and YC provided project administration and resources. ZW and YP wrote the original draft. JZ, YC, and SW supervised the study and performed writing – review & editing. All authors had full access to all the data in the study and had final responsibility for the decision to submit for publication. ZW and YP directly accessed and verified the underlying data reported in the manuscript.

## Acknowledgements

We are grateful for resources from the Bioinformatics Platform, Furong Laboratory and Bioinformatics Center, Xiangya Hospital, Central South University.

## Data Sharing Statement

The faers package is freely available through the Bioconductor repository at https://bioconductor.org/packages/faers/. Its source code is openly hosted on GitHub at https://github.com/WangLabCSU/faers. The raw FAERS data used for performance benchmarking and clinical validation are publicly accessible via the FDA’s Quarterly Data Files website (https://fis.fda.gov/extensions/FPD-QDE-FAERS/FPD-QDE-FAERS.html). To ensure reproducibility, all R scripts and source code used for the analyses in this manuscript are provided in the GitHub repository (https://github.com/MadDERt/faers-reproducibility).

